# Dimensionality reduction establishes specificity in lesion network mapping

**DOI:** 10.64898/2026.01.29.26345138

**Authors:** Shahar Edelman, Uri Elias, Shahar Arzy

**Affiliations:** Computational Neuropsychiatry Lab, Department of Medical Neurosciences, Faculty of Medicine, Hebrew University of Jerusalem, Jerusalem, Israel; Department of Neurology, Hadassah Hebrew University Medical School, Jerusalem, Israel; Department of Brain and Cognitive Sciences, Hebrew University of Jerusalem, Jerusalem, Israel

## Abstract

**Background:** Lesion network mapping (LNM) has emerged as a powerful tool for linking focal brain lesions to distributed functional networks. However, the biological specificity of these networks has been questioned. Recent mathematical derivations suggest that LNM-derived maps may trivially track the normative connectome’s global degree vector rather than specific symptom-related topography, potentially rendering them biologically nonspecific.

**Methods:** We introduced a rigorous validation pipeline to distinguish true network specificity from low-dimensional connectome artifacts. We projected lesion connectivity maps into a low-dimensional feature space defined by the principal gradients and eigenmodes of the normative connectome. We applied this framework to a large-scale dataset of 858 lesions associated with four distinct clinical cohorts: obsessive-compulsive disorder (OCD), schizophrenia, aphasia, and epilepsy. We performed multivariate classification to determine if symptom-associated lesions occupied distinct regions of the functional manifold compared to null distributions.

**Results:** Our analysis revealed a sharp dissociation in network specificity across disorders. While schizophrenia-associated lesions were indistinguishable from null models (Accuracy=0.51, p=0.412), confirming the “degree artifact” hypothesis for this cohort, other disorders displayed significant network specificity. Lesions associated with OCD (Accuracy=0.58, p=0.036), aphasia (Accuracy=0.60, p=0.007), and epilepsy (Accuracy=0.61, p=0.002) occupied distinct regions of the functional manifold significantly different from the normative connectome baseline.

**Conclusions:** These findings demonstrate that while LNM is sensitive to connectome-level artifacts, it retains genuine biological specificity for distinct clinical phenotypes. The proposed linear projection framework offers a standardized, computationally efficient benchmark for assessing network specificity against methodological noise.

## Introduction

Lesion network mapping (LNM) is a fascinating method, developed by Michael D. Fox and colleagues, to address a longstanding puzzle in cognitive neurology: how anatomically distinct brain lesions can underlie highly similar clinical symptoms. Instead of assuming a disrupted, localized module, LNM adopts a network-based perspective, positing that disparate lesions may converge on a common functional network subserving the affected cognitive function.^1^ In practice, the method is as follows: each lesion is treated as a seed region, and then, using large-scale normative connectivity data, the distributed network functionally connected to that lesion is derived; these networks are thresholded, and their overlap is defined as the symptom-related network.^1,2^

A recent work by van den Heuvel et al.^3^ exposed a “foundational limitation” in LNM, demonstrating that the method is effectively a linear operation (*LNM* ≈ *M* × C, where *M* rows are lesions and *C* is a derivation of the normative connectome). It had been argued that since LNM-derived maps are effectively sampling the rows of *C*, they often converge to the elementary properties of *C* - specifically its row-summation vector or degree. Consequently, resulting networks may be largely nonspecific and biologically nondistinct. This critique highlights an important point, namely, that simply averaging and thresholding functional connectivity maps may create an artifact of convergence to the degree vector of *C*.

However, the mathematical formalization provided by van den Heuvel et al. does not inherently invalidate the LNM framework but rather provides the necessary tools to empirically test its specificity. Van den Heuvel et. al. noted that 93% of the variance in reported LNM networks is explained by basic properties of the normative connectome. While this has been interpreted as evidence for lack of specificity, we argue that this may reflect the idea of biological data compression that is found in the basis of the LNM paradigm: the premise that heterogenous lesion effects are constrained to a low-dimensional manifold defined by brain connectivity, and that these connectome-based maps might explain symptoms in a way that discrete anatomical lesion analyses cannot.^1,2^ The shown “flaw” in LNM is not the low dimensionality of its outputs, but the statistical approach of averaging high-dimensional data into a single map that inevitably collapses toward the row summation of the matrix *C*.

## Methods

To address this, we implemented a validation pipeline that abandons voxel-wise averaging in favor of a litmus test exploiting the linear approximation (*M* × *C*).

Foremost, the low dimensionality of the process enables direct statistical analysis against behavioral data, resolving the common statistical imbalance between the high dimensionality of the brain and the typically limited number of patient samples. Furthermore, the linear approximation reduces computation time for a standard dataset from hours to under 10 seconds. This efficiency enables the generation of large-scale null lesion distributions that were previously computationally prohibitive.

Our validation pipeline proceeded as follows:

1. **Generate null models:** We constructed a matrix *M* ∈ *R*^*P*×*d*^ (Where *P* is number of lesions and *d* is dimension of the parcellation), where rows represent null lesions drawn from large empirical datasets. We used the linear approximation (*M* × *C*) to efficiently generate functional connectivity maps for this null distribution, and performed the identical operation for the symptom-positive patient cohort.
2. **Projection**: Instead of analyzing voxel-wise overlap, we projected both symptom-positive and symptom-negative connectivity maps into the low-dimensional subspace defined by the properties of *C*.
3. **Individual-level predictions**: Within this subspace, we performed multivariate classification (k-nearest neighbors) on individual maps.

This approach tests patient lesions will be separable from the random null distribution in this low-dimensional space. By sharing lesion and behavioral data openly, the community can populate this null space and to transparently see the effectiveness of network mapping, turning this critique into a standardized benchmark for establishing genuine network phenotypes.

## Results

We conducted this analysis using open-source data available in the van den Heuvel et al. repository. We projected individual lesion connectivity maps (computed via the linear approximation) into the low-dimensional feature space described in the original critique (comprising global connectivity degree, subcortical connectivity, resting-state network module strengths, and the first three principal gradients of *C*), yielding a 9-dimensional representation per lesion.

We analyzed four datasets including more than 100 subjects: OCD^4^, schizophrenia^5^, aphasia recovery,^6^ epilepsy.^7^ For each dataset, we performed a binary classification using k-nearest neighbors with nested cross-validation (5-fold outer, 3-fold inner to tune K), against a similar number of negative lesions that were randomly sampled from the other lesion sets (*n* = 858 total lesions). We assessed statistical significance against the null distribution using permutation testing (1000 permutations).

Crucially, this individual-level approach disentangled cases that appeared identical under the averaging paradigm. Van den Heuvel et. al. noted that averaged maps for both OCD and schizophrenia (*n* = 100 each) were nearly indistinguishable from the connectome’s degree vector (*r* = 0.96, 0.97respectively), implying LNM failure for both. However, our classification revealed a sharp distinction: while we could not significantly separate schizophrenia-related lesions from the null distribution (*Accuracy* = 0.51, p = 0.412), we achieved significant separability for OCD (*Accuracy* = 0.58, p = 0.036). We similarly found significant classification accuracy for aphasia recovery (*Accuracy* = 0.60, p = 0.007) and epilepsy (*Accuracy* = 0.61, p = 0.002).

## Discussion

These results suggest that certain disorders occupy genuinely distinct regions of the functional connectivity manifold, supporting the view that lesion connectivity may serve as a valuable framework to link heterogeneous lesions into a pattern in the functional connectivity space. While this demonstrates the utility of LNM for specific phenotypes, its application to other disorders requires rigorous re-examination. Furthermore, even where separability is demonstrated, the validity of existing published maps remains questionable; as products of averaging, these maps likely reflect the connectome’s degree centrality rather than a true “network localization of neurological symptoms”.^8^

While LNM may require methodological corrections, it would be a mistake to reject the prominent paradigm of using the human connectome as a possible bridge between anatomical heterogeneity and shared phenomenology. Seed-based connectivity has successfully recapitulated canonical networks (e.g.,^9^) and provides a vital tool for linking neurology to behavior. The approximation of LNM using linear projection into a low-dimensional subspace does not invalidate the field; rather, it creates an opportunity to develop connectivity techniques with unprecedented efficiency. Critically, the bottleneck of small sample sizes is no longer a prohibitive limitation: the computational speed enables creating larger datasets seamlessly while the low dimension ensures robust statistical power. Our proposed pipeline represents just one such avenue. Future work can now leverage this low-dimensional framework to explore broader questions, such as whether clustering within the connectivity manifold aligns with shared clinical properties.

According to the law of the instrument, (Maslow’s hammer) it is tempting, if the only tool you have is a hammer, to treat everything as if it were a nail.^10,11^ While this bias is indeed true, a hammer is nonetheless a much needed tool.

## Data Availability

Parcellation pipelines, the normative connectome matrix (C), original data sources, and other useful scripts were adopted from the van den Heuvel et al. (2026) GitHub repository at https://github.com/dutchconnectomelab/lesionnetworkmapping. All scripts and data used to create the results in this work are available at https://codeocean.com/capsule/8846447/.

